# Clinical outcomes of flomoxef versus cefmetazole in hospitalized patients with urinary tract infections: Combined retrospective analyses of two real-world databases and *in vitro* data

**DOI:** 10.1101/2024.07.16.24310469

**Authors:** Takahiro Niimura, Mitsuhiro Goda, Satoshi Nakano, Toshiki Kajihara, Koji Yahara, Aki Hirabayashi, Koji Miyata, Marie Ikai, Motoko Shinohara, Yusuke Minato, Masato Suzuki, Keisuke Ishizawa

**Affiliations:** Department of Clinical Pharmacology and Therapeutics, University of Tokushima Graduate School of Biomedical Sciences, Tokushima, Japan; Clinical Research Center for Developmental Therapeutics, Tokushima University Hospital, Tokushima, 770-8503, Japan; Department of Pharmacy, Tokushima University Hospital, Tokushima, Japan; Antimicrobial Resistance Research Center, National Institute of Infectious Diseases, Tokyo, Japan; Center for Infectious Disease Research, Fujita Health University, Aichi, Japan

**Keywords:** cefmetazole, flomoxef, urinary tract infection

## Abstract

**Background:** Flomoxef is active against extended-spectrum beta-lactamase (ESBL)-producing bacteria, but its clinical effectiveness has not been compared with cefmetazole. This study aimed to compare the utility of the two drugs in treating urinary tract infection (UTI) by integrating *in vitro* data with a retrospective analysis of two real-world databases.

**Methods:** The susceptibility rates of third-generation cephalosporin-resistant *Escherichia coli* and *Klebsiella pneumoniae* to flomoxef and cefmetazole were compared using comprehensive national antimicrobial resistance surveillance data. Combinational antimicrobial activities against an ESBL-producing multidrug-resistant bacterial strain were tested by diagonal measurement of n-way drug interactions (DiaMOND), an innovative method to assess antimicrobial interactions *in vitro*. The effectiveness of the two drugs in treating UTIs was compared using hospital stay duration as the primary outcome obtained from the Japan Medical Data Center (JMDC) Claims Database.

**Results:** Third-generation cephalosporin-resistant *E. coli* and *K. pneumoniae*, including ESBL-producing strains, were similarly susceptible to flomoxef and cefmetazole. *In vitro* assessment of combinational antimicrobial activities against an ESBL-producing multidrug-resistant strain revealed that both drugs showed similar antimicrobial interaction patterns.

JMDC Claims data analysis showed that the median time of hospital stay duration was 11 days (95% confidence interval [CI]: 11–11) in the cefmetazole group and 4 days (95% CI: 3– 5) in the flomoxef group, significantly shorter in the latter (log-rank test, P < 0.001). In addition, the flomoxef group had a significantly lower frequency of adverse events such as *Clostridioides difficile* infection.

**Conclusions:** Flomoxef exhibits effectiveness that is comparable to cefmetazole in treating UTI without major concerns about adverse events such as *C. difficile* infection or renal impairment. This evidence endorses flomoxef as a viable treatment option for UTIs in locales with high prevalence of ESBL-prodcing strains.

**Key Points:** An oxacephem antimicrobial agent, flomoxef, exhibited effectiveness comparable to cefmetazole in the treatment of urinary tract infections and was superior in terms of safety.

## Introduction

The increase in extended-spectrum beta-lactamase (ESBL)-producing bacteria has complicated the treatment of urinary tract infections (UTI) and intra-abdominal infections due to resistance to various classes of antibiotics [1–5]. Currently, carbapenems are the preferred treatment option for severe infections caused by ESBL-producing bacteria. On the other hand, carbapenem-resistant strains have emerged and spread, which has raised interest in treatment strategies that spare carbapenems [6].

Cefmetazole and flomoxef are antimicrobial agents originally developed in Japan. Cefmetazole is a cephamycin that has a broad spectrum of antibacterial activity against *Enterobacterales*, such as *Escherichia coli* and *Klebsiella pneumoniae,* as well as anaerobic bacteria and is widely indicated for the treatment of UTI and intra-abdominal infections [7]. Flomoxef is an oxacephem that, like cefmetazole, is active against *E. coli*, *K. pneumoniae* and other *Enterobacterales*, and is indicated for UTI and intra-abdominal infections [8, 9]. Both cefmetazole and flomoxef have a methoxy group at the 7-position of cephalosporanic acid and are stable to hydrolysis by ESBL [10]. Cefmetazole and flomoxef demonstrate *in vitro* efficacy against ESBL-producing bacteria and are of increasing clinical importance as carbapenem-sparing therapeutic options for infections caused by ESBL-producing *Enterobacterales* [11, 12].

Although clinical data are accumulating that suggest comparable effectiveness of cefmetazole with carbapenems against infections due to ESBL-producing bacteria, clinical data regarding flomoxef remain limited [13, 14]. To the best of our knowledge, there has been no national study comparing the susceptibility rates of flomoxef and cefmetazole among ESBL-producing *Enterobacterales*. Furthermore, there has been no large study comparing the effectiveness of treatment with flomoxef and cefmetazole. Considering the unstable supply of antimicrobials in recent years [15, 16], it would be beneficial to clarify the potential of flomoxef as a treatment option against ESBL-producing bacterial infections.

This study aimed to compare the usefulness of the two drugs by integrating *in vitro* data with a retrospective analysis of two real-world databases (National claims database and Nosocomial infections surveillance database).

## Methods

### Antimicrobial susceptibility profile of *E. coli* and *K. pneumoniae* clinical isolates based on national antimicrobial resistance surveillance data

Data on bacterial isolates from inpatients in Japan between January 2014 and December 2021 were extracted from the national antimicrobial resistance surveillance program, the Japan Nosocomial Infections Surveillance (JANIS), which collects all routine microbiological diagnostic tests [17]. A total of 2,220 hospitals across Japan submitted their data to the JANIS database in 2021. We specifically targeted *E. coli* and *K. pneumoniae* due to their prevalence in UTIs. Data on antimicrobial susceptibility testing of isolates from urine and blood specimens was obtained, and the MIC values of cefotaxime, cefmetazole, and flomoxef were analyzed. We interpreted the MIC data of cefotaxime and cefmetazole using the Clinical and Laboratory Standards Institute (CLSI) 2023 criteria [18], which have not changed since the CLSI 2012. In this study, cefotaxime-resistant *E. coli* and *K. pneumoniae* isolates were considered as third-generation cefphalosporin-resistant isolates.

The part of this study was approved by the Ministry of Health, Labour and Welfare (approval number: 1025–2) according to Article 32 of the Statistics Act.

### *In vitro* drug interaction characterization

Drug interactions were assessed using the DiaMOND method as previously described [19, 20]. An ESBL-producing, multidrug-resistant uropathogenic *E. coli* clinical strain UPEC GU2019-E4 [21] was used. Unless otherwise noted, bacterial cells were grown in LB medium at 37°C under aerobic conditions. The bacterial cultures were diluted in fresh medium to an OD_600_ of 0.001, and 50 μL of the diluted culture was added to each well of a clear flat-bottom 384-well microplate. Antimicrobial agents were dispensed using a digital dispenser (Multidrop pico8 digital dispenser; Thermo Fisher). After 22 hrs incubation at 37°C without shaking, OD_600_ was measured using a microplate reader (Bio Tek Synergy HTX plate reader: Agilent). The fractional inhibitory concentration index (FICI) was used to evaluate the combination effect of the drugs. Dose-response sampling for the combination was performed by measuring and calculating the IC_90_ values for each drug separately and creating a dilution series of the combined drugs at half the concentration of each drug’s IC_90_ values. The growth inhibition assay was performed as with the single drugs, and the OD_600_ data obtained were used to calculate the IC_90_, which was defined as the observed IC_90_. The intersection of the line connecting the IC_90_ values of each single drug and the diagonal line representing the dose-response of the combination drug was defined as the expected IC_90_.

## Data source of claims database analysis

Data from April 2014 to November 2021 in the JMDC Claims Database were used for the study. This database contains inpatient and outpatient claims, as well as Diagnosis Procedure Combination (DPC) practice data from several health insurance associations [22]. The DPC system is a comprehensive reimbursement system for acute inpatient care introduced in Japan in 2003 [23]. From the insurance claims data and DPC practice data, it is possible to extract diagnoses of injuries and diseases, drug prescriptions, weight, height, and activities of daily living (ADL) scores at the time of admission.

This part of the study was approved by the Ethics Committee of Tokushima University Hospital, protocol number: 4492. This study was conducted in accordance with the Strengthening the Reporting of Observational Studies in Epidemiology (STROBE) Reporting Guidelines for Cohort Studies.

## Study design of claims database analysis

Patients with a confirmed diagnosis of UTI on the date of first admission after records began to be collected from each patient’s JMDC and who were prescribed cefmetazole or flomoxef on the same day were included. The diagnosis of UTI was based on the International Statistical Classification of Diseases and Related Health Problems 10th Revision (ICD-10) Version 2016 of the World Health Organization (Supplementary Table 1). The date of the first cefmetazole or flomoxef dose was set as the index date (Fig. 1). Certain patient groups were excluded: those not in the DPC wards, those without a UTI diagnosis on the antimicrobial prescription date, those who died within 24 hours of admission, and those receiving both cefmetazole and flomoxef.

**Figure 1.**
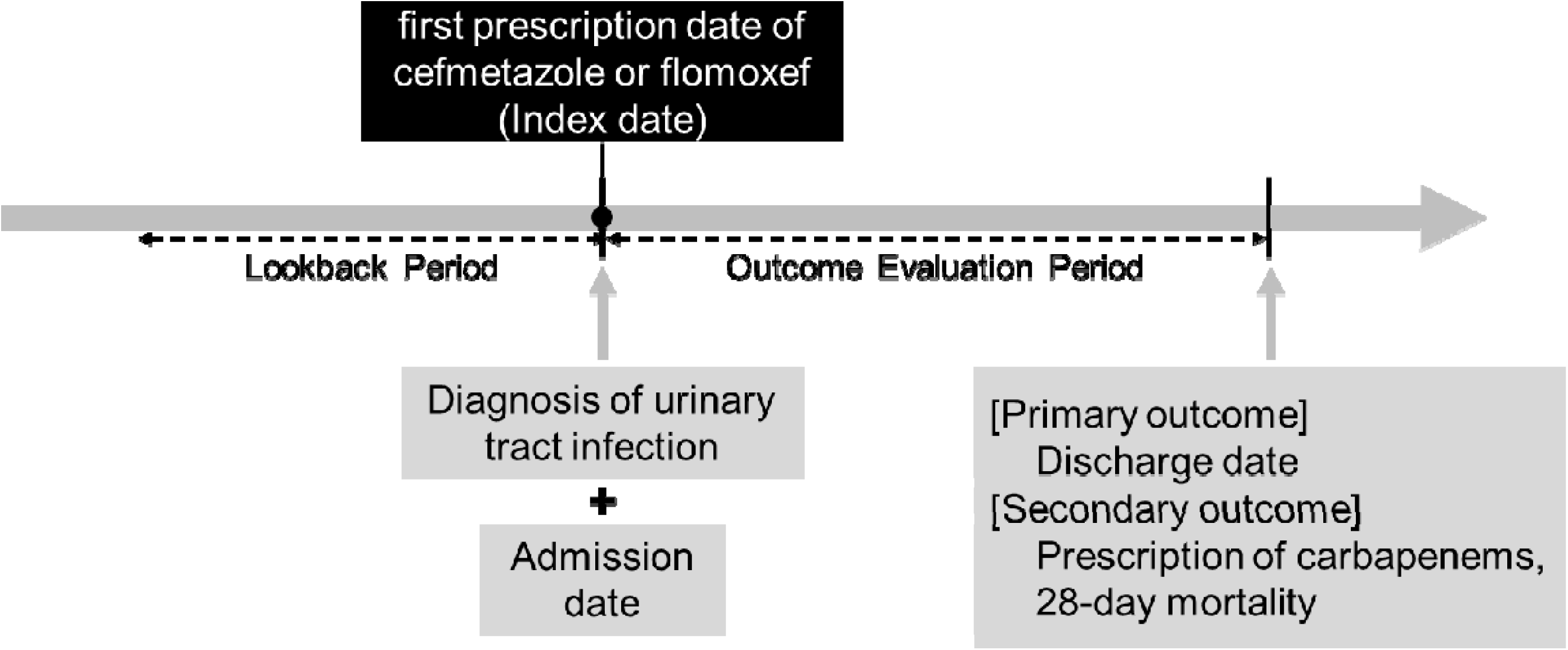
Analysis design of claim data analysis. Time line for index date and associated time point. Abbreviations: JMDC, Japan Medical Data Center.

Because of the multiple outcomes and statistical methods used in this study, several analytic datasets were created: Dataset 1 was the dataset of all remaining patients after excluding those who met the exclusion criteria; Dataset 2 consisted of only those patients who were discharged alive after excluding those who died during hospitalization; Dataset 3 was created by 1:1 matching of patients given cefmetazole and flomoxef by the propensity score for Dataset 2.

## Definition of baseline characteristics and complications

Sex, age, body mass index (BMI), Barthel index, Japan coma scale (JCS), and various complications (sepsis, shock, respiratory failure) were evaluated at the index date. BMI was classified as underweight (BMI <18.5 kg/m^2^), normal weight (BMI 18.5–24.9 kg/m^2^), overweight (BMI 25.0–29.9 kg/m^2^), obese (BMI ≥30 kg/m^2^), and deficient, according to previous studies [24]. The Barthel Index consists of 10 items: feeding (0–10 points), bathing (0–5 points), grooming (0–5 points), dressing (0–10 points), bowel control (0–10 points), bladder control (0–10 points), voiding (0–10 points), chair movement (0–15 points), walking (0–15 points), and stair climbing (0–10 points). A higher score indicated a higher degree of independence in ADLs. The JCS categorizes consciousness into four major grades based on reactive eye-opening. Grade 0 signifies awake consciousness. Grades 1–3, represented by a single digit, indicate awake status without any stimulus. Grades 10–30, denoted by two digits, describe a state where the individual awakens with stimulus but reverts to the previous state once the stimulus is removed. Finally, grades 100–300, expressed with three digits, correspond to a state where the individual does not arouse in response to any stimulus [25]. The JCS has been reported to correlate well with the globally used Glasgow coma scale [26]. The complications of sepsis, shock, and respiratory failure were evaluated based on the diagnoses listed in Supplementary Table 2. The Charlson comorbidity index and history of diabetes were evaluated based on diagnoses before index date (Fig. 1). The Charlson comobidity index was assigned to each patient according to a previous study [27]. History of diabetes or urinary tract abnormalities was defined based on the ICD-10 codes (Supplementary Table 3).

## Patient outcomes

The primary effectiveness outcome was the number of days from the index date to discharge. Secondary effectiveness outcomes were defined as 28-day mortality after the index date and carbapenem use within 7 days after the index date. Safety outcomes were defined as gastrointestinal adverse events (AEs) (non-*C. difficile* diarrhea, *C. difficile* infection), renal-related AEs (kidney failure and other kidney dysfunctions), and hypersensitivity symptoms (anaphylactic shock and rash) (Supplementary Table 4). Gastrointestinal– and renal-related AEs were analyzed one month after the index date, and hypersensitivity symptoms were analyzed two days after the index date. AEs were defined based on the ICD-10 codes (Supplementary Table 4).

The primary effectiveness outcome was analyzed using Datasets 2 and 3, and the secondary effectiveness outcomes were analyzed using Dataset 1. Safety outcomes were analyzed using Dataset 1.

## Statistical analyses

Continuous variables were expressed as mean ± standard deviation, while categorical variables were reported as percentages. The standardized mean difference was used to identify differences in patient backgrounds between the cefmetazole and flomoxef groups. Kaplan–Meier analysis was used to estimate the primary effectiveness outcome, namely the time from the initiation of cefmetazole or flomoxef treatment to discharge alive, and log-rank tests were used to evaluate differences. Cox regression analysis was performed using age, sex, Charlson comorbidity index, BMI, JCS, complications (sepsis, shock, respiratory failure), and underlying urinary tract abnormalities as covariates that may affect prognosis. Hazard ratios (HRs) for survival and discharge were calculated. Propensity score matching with 1:1 matching was performed to balance baseline characteristics. The propensity score was calculated using age, sex, Charlson comorbidity index, BMI, JCS, comorbidities (sepsis, shock, respiratory failure), underlying urinary tract abnormalities, and Barthel index. Kaplan– Meier analysis, log-rank test, and Cox regression analysis were performed on the crude and matched data.

For the secondary effectiveness outcome, logistic regression analysis was performed with age, sex, Charlson Comorbidity Index, BMI, JCS, complications (sepsis, shock, respiratory failure), and underlying urinary tract abnormalities as covariates. The odds ratios for 28-day mortality and prescriptions of carbapenems were calculated.

For safety outcomes, the frequencies were calculated and descriptively analyzed for gastrointestinal, renal, hepatic, and hypersensitive AEs.

Statistical tests were two-tailed, with a p-value of less than 0.05 considered statistically significant, and performed using R version 4.2.1 (R Foundation for Statistical Computing, Vienna, Austria).

## Results

### Antimicrobial susceptibility of *E. coli* and *K. pneumoniae* isolated from urine and blood against cefmetazole and flomoxef

In total, 682,034 *E. coli* and 143,842 *K. pneumoniae* isolates from urine culture specimens, and 144,219 *E. coli* and 53,334 *K. pneumoniae* isolates from blood culture specimens underwent antimicrobial susceptibility testing for both cefmetazole and flomoxef. Among them, 1.4% of 3rd generation cephalosporin-resistant (3GC-R) *E. coli* isolates and 5.5% of 3GC-R *K. pneumoniae* isolates from urine had MICs of cefmetazole of ≥64 mg/L. On the other hand, 1.0% of 3GC-R *E. coli* isolates and 3.0 % of 3GC-R *K. pneumoniae* isolates had MICs of flomoxef of ≥64 mg/L. Concerning blood culture specimens, 1.5% of 3GC-R *E. coli* isolates and 6.2% of 3GC-R *K. pneumoniae* isolates had MICs of cefmetazole of ≥64 mg/L. Meanwhile, 1.1% of 3GC-R *E. coli* isolates and 2.8 % of 3GC-R *K. pneumoniae* isolates had MICs of flomoxef of ≥64 mg/L.

### Interaction of cefmetazole and flomoxef with other classes of antimicrobial agents

We next compared antimicrobial drug interactions of cefmetazole and flomoxef against an ESBL-producing, multidrug-resistant *E. coli* strain. The checkerboard assay is the standard method to determine FICI as a measure of interaction between antimicrobial agents; however, the assay is labor-intensive and time-consuming. Thus, we employed the DiaMOND (diagonal measurement of n-way drug interactions), a recently developed high-throughput method to assess antimicrobial interactions *in vitro* (Fig. 2). As shown in Fig. 2A, we first determined IC_50_ and IC_90_ values of cefmetazole, flomoxef, and ten other antimicrobial drugs against *E. coli* GU2019-E4 (Fig. 2B). We then compared antimicrobial drug interactions of cefmetazole and flomoxef with ten different antimicrobial agents (Fig. 2C). We found that the two drugs showed similar antimicrobial drug interaction patterns, supporting our hypothesis that flomoxef is as active as cefmetazole, both by itself and in combination with other commonly used agents.

**Figure 2.**
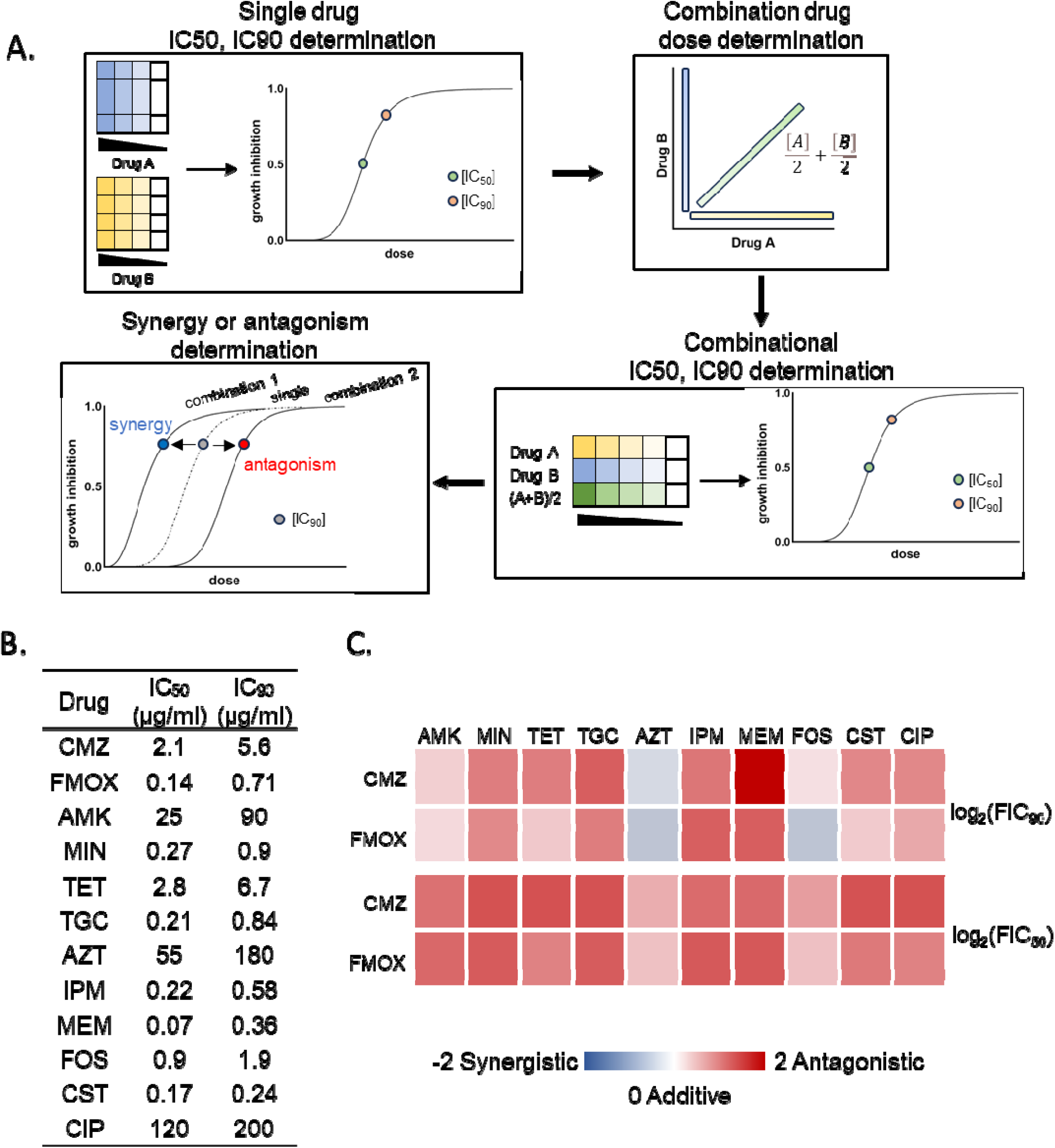
antibacterial drug combination effects. (A) Experimental and analysis workflow for measuring drug combination effects using DiaMOND method. (B) IC_50_ and IC_90_ of the antibacterial drugs against E.coli GU2019-E4 strain. CMZ, cefmetazole; FMOX, flomoxef; AMK, amikacin; MIN, minocycline; TET, tetracycline; TGC, tigecycline; AZT, aztreonam; IPM, imipenem; MEM, meropenem; FOS, fosfomycin; CST, collistin; CIP, ciprofloxacin. GEN, gentamicin; STR, streptomycin; TOB, tobramycin; (C) Heatmap of the combination effects of 10 antimicrobial drugs on CMZ and FMOX. Drug interactions are assessed using log2(FIC_50_) and log2(FIC_90_) values: a log2(FIC) below 0 indicates synergy (represented in blue) and a log2(FIC) above 0 indicates antagonism (represented in red).

### Baseline characteristics of JMDC Claims Database

Between April 2014 and November 2021, 340,954 and 83,932 patients were prescribed cefmetazole and flomoxef in the JMDC database, respectively. The study comprised 4796 patients who received cefmetazole and 673 patients who received flomoxef (Fig. 3). This group formed Dataset 1. Dataset 2, which excluded patients who died in hospital from Dataset 1, comprised 4552 and 658 patients treated with cefmetazole and flomoxef, respectively. In Datasets 1 and 2, before propensity score matching, there were significant differences between the two groups regarding age, sex, BMI, Barthel index, history of diabetes, complications (sepsis, shock, respiratory failure), and underlying urinary tract abnormalities (Table 2). Patients who received cefmetazole were more likely to be female, older, and have history of diabetes mellitus and complications, such as sepsis, shock, and respiratory failure than those who received flomoxef. Moreover, based on the JCS score, the state of consciousness was worse in the cefmetazole group than in the flomoxef group. In contrast, the flomoxef group had a higher proportion of patients with underlying urinary tract abnormalities than the cefmetazole group. After matching by propensity score, the two groups were similar with respect to baseline characteristics, except for history of diabetes.

**Figure 3.**
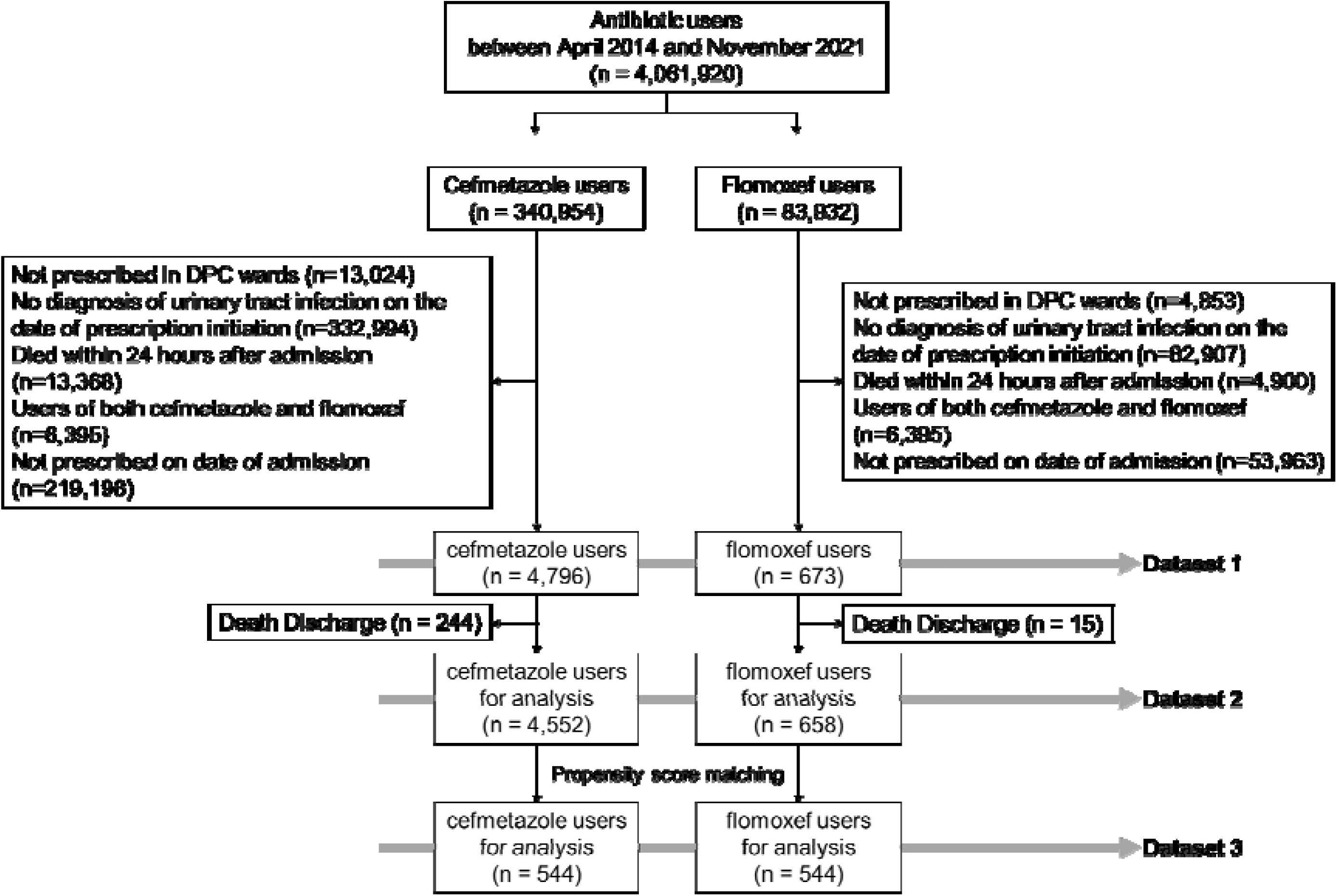
Patient flowchart in the claim data analysis. Flow diagram of patient inclusion and set up of each dataset. Abbreviations: DPC, Diagnosis Procedure Combination.

**Table 1.** The MIC values for cefmetazole and flomoxef against *E. coli* and *K. pneumoniae* isolated from urine and blood samples.

| Specimen | Isolates | Total | Cefmetazole |  |  | Flomoxef |  |  |
| --- | --- | --- | --- | --- | --- | --- | --- | --- |
|  |  |  | ≤16 | 32 | ≥64 | ≤16 | 32 | ≥64 |
| Urine | *E. coli* (all) | 682,034 | 674,654 (98.9) | 4,240 (0.6) | 3,140 (0.5) | 675,602 (99.0) | 4,476 (0.7) | 1,956 (0.3) |
|  | 3rd Generation cephalosporin-resistant<br>*E. coli* | 164,276 | 159,774 (97.3) | 2,180 (1.3) | 2,322 (1.4) | 160,129 (97.5) | 2,489 (1.5) | 1,658 (1.0) |
|  | *K. pneumoniae* (all) | 143,842 | 141,938 (98.7) | 649 (0.5) | 1,255 (0.9) | 142,537 (99.1) | 744 (0.5) | 561 (0.4) |
|  | 3rd Generation cephalosporin-resistant<br>*K. pneumoniae* | 14,937 | 13,843 (92.7) | 272 (1.8) | 822 (5.5) | 14,051 (94.1) | 439 (2.9) | 447 (3.0) |
| Blood | *E. coli* (all) | 144,219 | 142,719 (99.0) | 872 (0.6) | 628 (0.4) | 142,964 (99.1) | 853 (0.6) | 402 (0.3) |
|  | 3rd Generation cephalosporin-resistant<br>*E. coli* | 31,455 | 30,516 (97.0) | 468 (1.5) | 471 (1.5) | 30,606 (97.3) | 493 (1.6) | 356 (1.1) |
|  | *K. pneumoniae* (all) | 53,334 | 52,661 (98.7) | 254 (0.5) | 419 (0.8) | 52,949 (99.3) | 247 (0.5) | 138 (0.2) |
|  | 3rd Generation cephalosporin-resistant<br>*K. pneumoniae* | 4,053 | 3,701 (91.3) | 101 (2.5) | 251 (6.2) | 3,797 (93.7) | 141 (3.5) | 115 (2.8) |

**Table 2.**
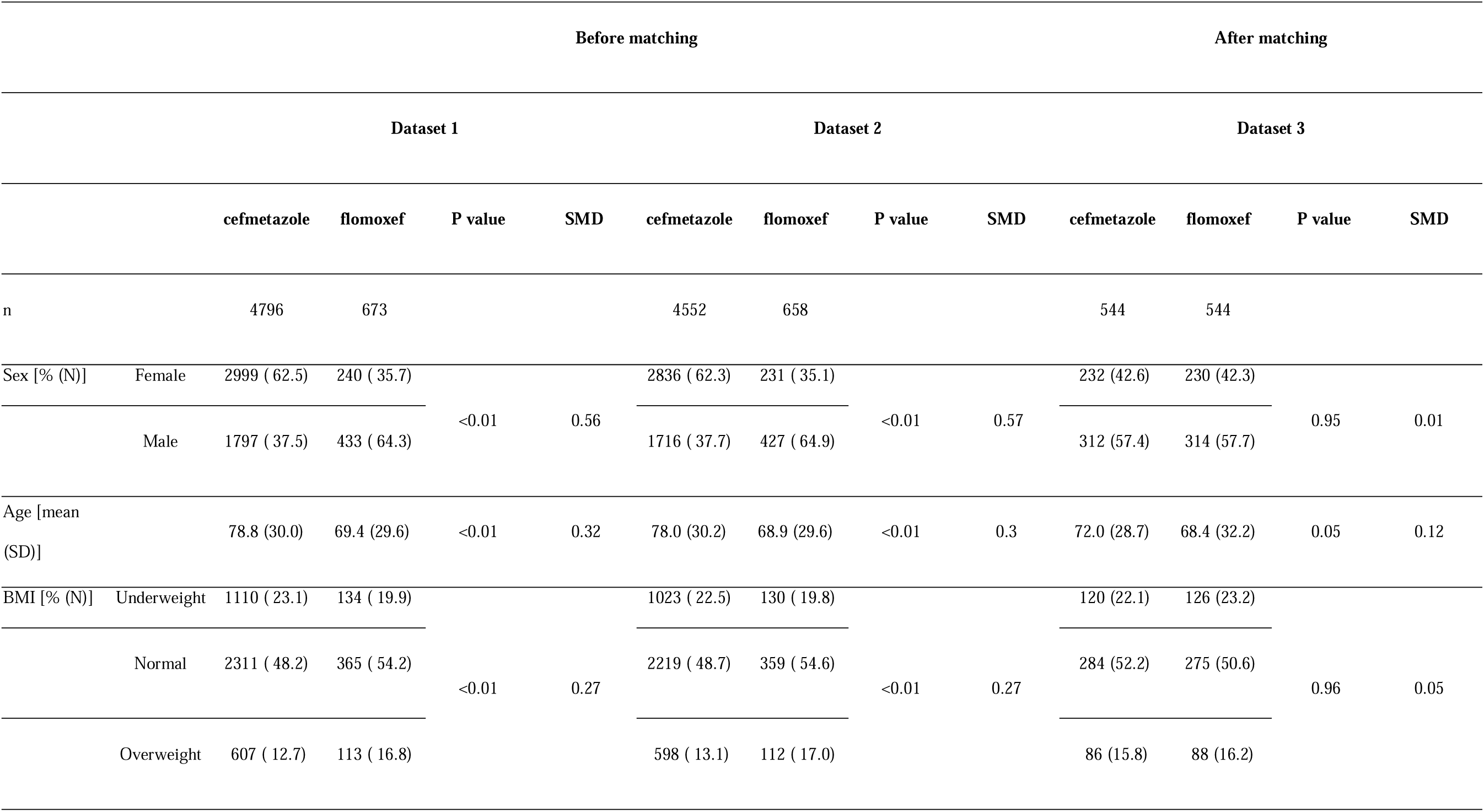
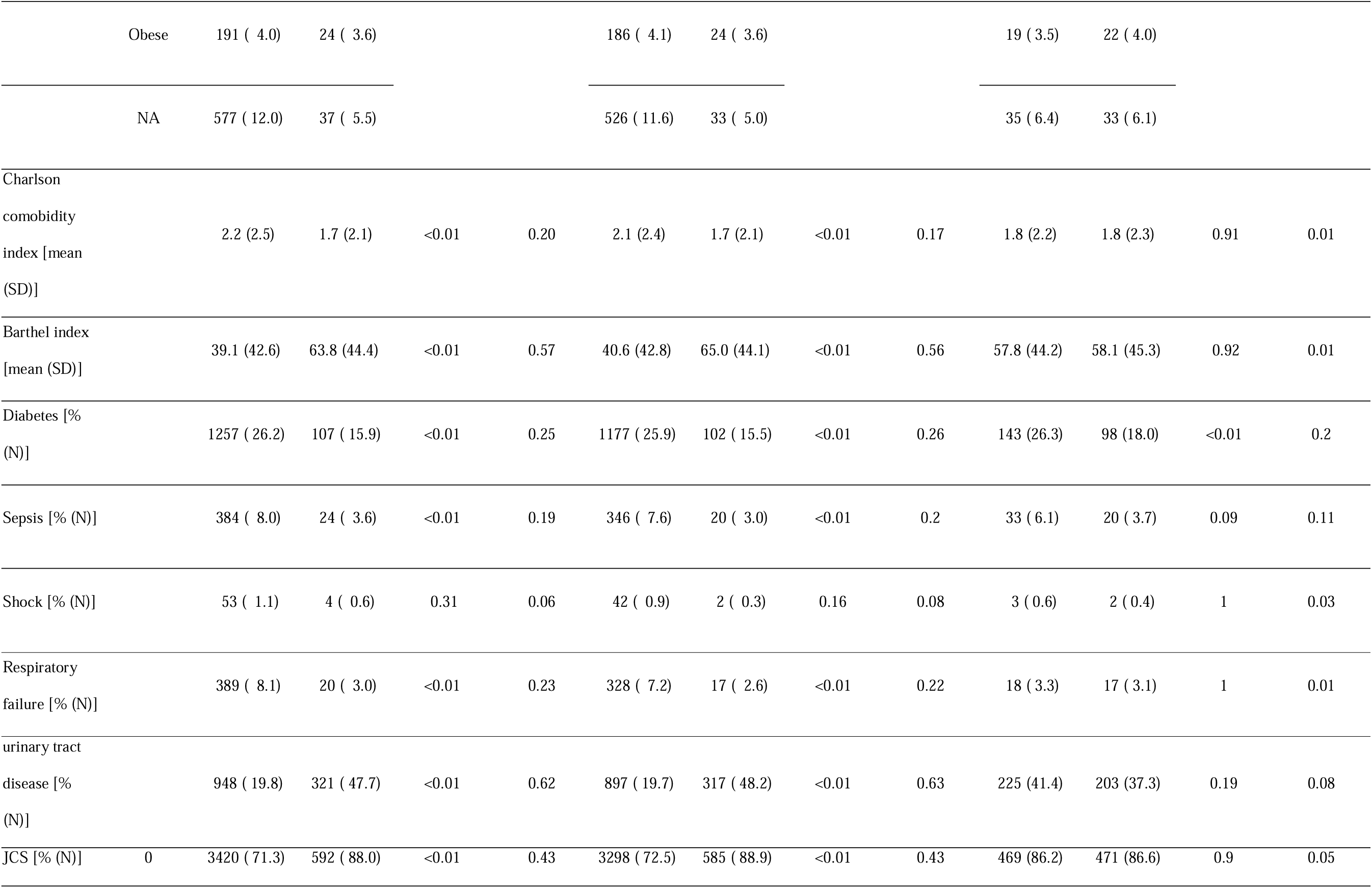
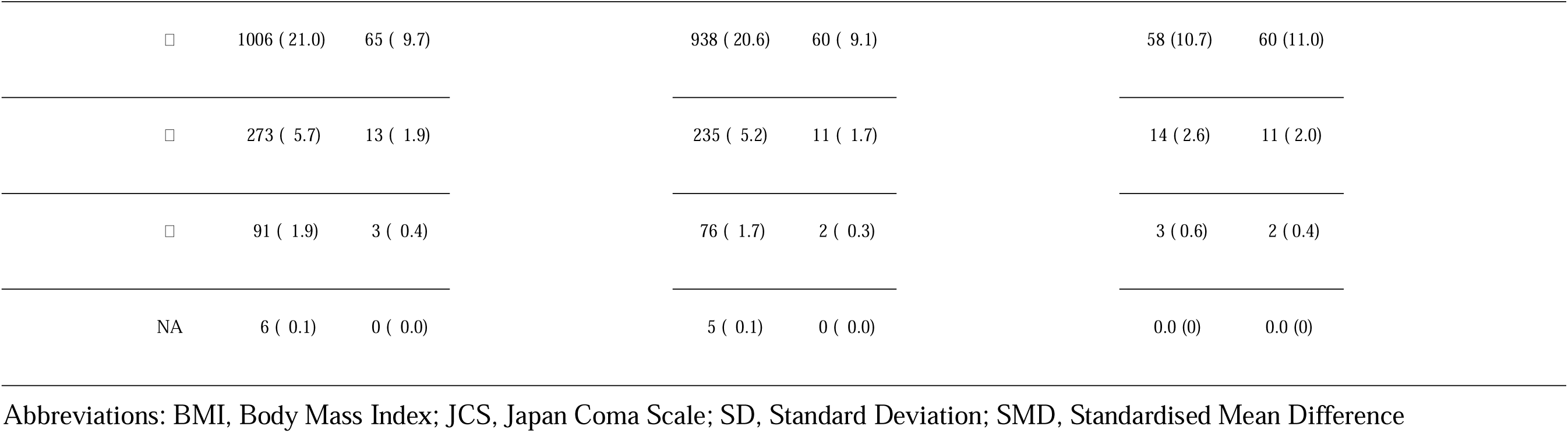
Characteristics of the Patients at Baseline.

### Primary effectiveness outcome

Dataset 2 was used to compare the two groups in terms of days to hospital discharge. The median time to discharge was 11 days (95% Confidence Interval [CI]: 11–11) in the cefmetazole group and 4 days (95% CI: 3–5) in the flomoxef group, with significantly fewer days in the flomoxef group (log-rank test: P<0.001) (Fig. 4). Cox proportional hazards analysis also revealed an HR of 1.75 (95% CI: 1.61–1.90, P<0.001) for live discharge in the flomoxef group.

**Figure 4.**
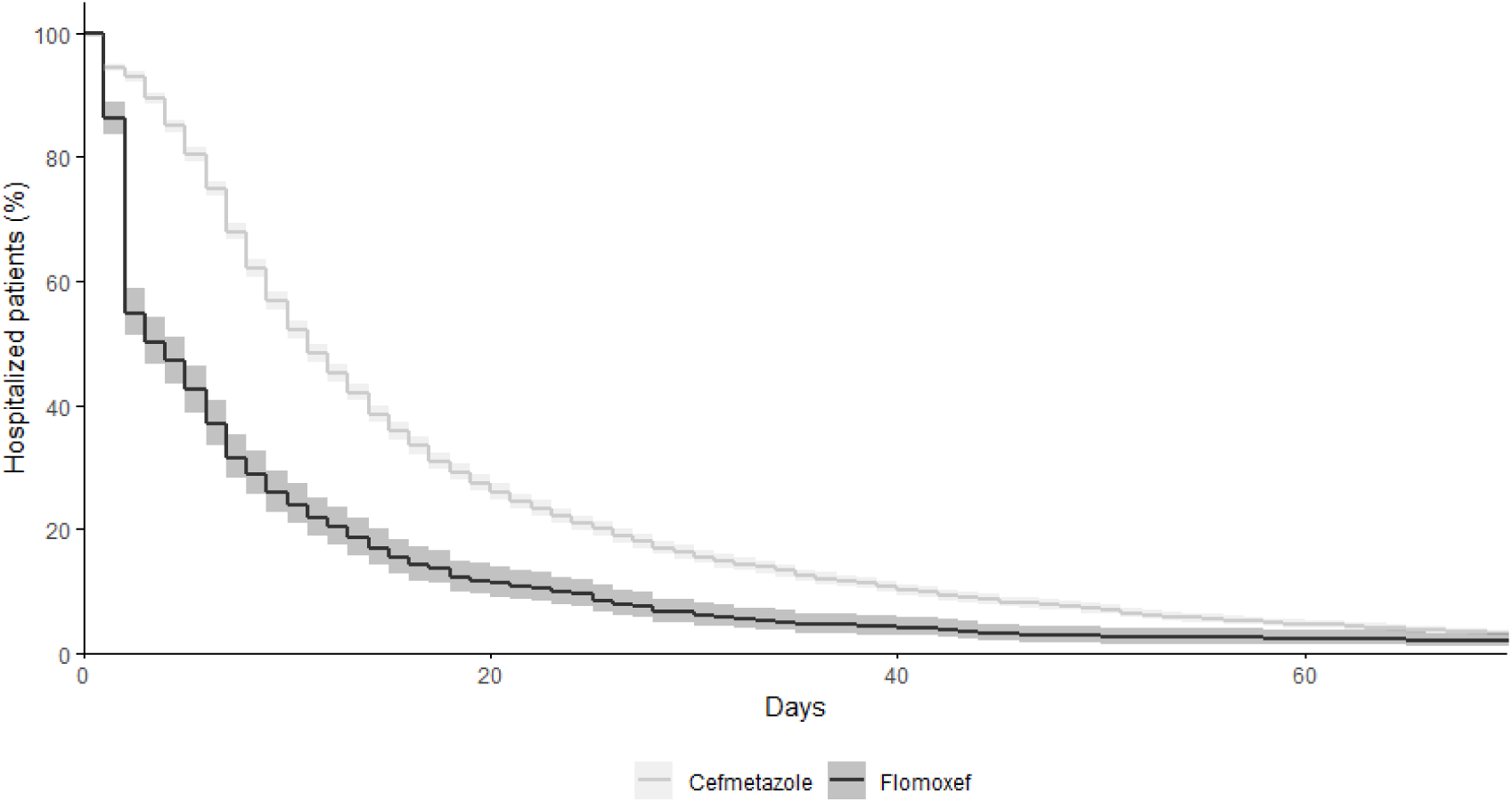
Kaplan–Meier plot illustrating survival post-discharge in Dataset 2. Kaplan-Meier curves for length of hospital stay by antimicrobial agent. Kaplan-Meier plots show hospitalization rates for patients in the cefmetazole and flomoxef groups from index date to survival discharge for Dataset 2.

The duration of hospital stay until discharge was evaluated using Dataset 3, following propensity score matching. In this analysis, the median time to discharge was 8.5 days (95% CI: 8–9) for patients in the cefmetazole group, compared to 5 days (95% CI: 5–6) for those in the flomoxef group. Thus, the flomoxef group experienced significantly shorter hospital stays, as evidenced even after propensity score matching (log-rank test: P<0.001) (Fig. 5). The hazard ratio for live discharge associated with flomoxef use was 1.60 (95% CI: 1.41–1.82, P<0.001).

**Figure 5.**
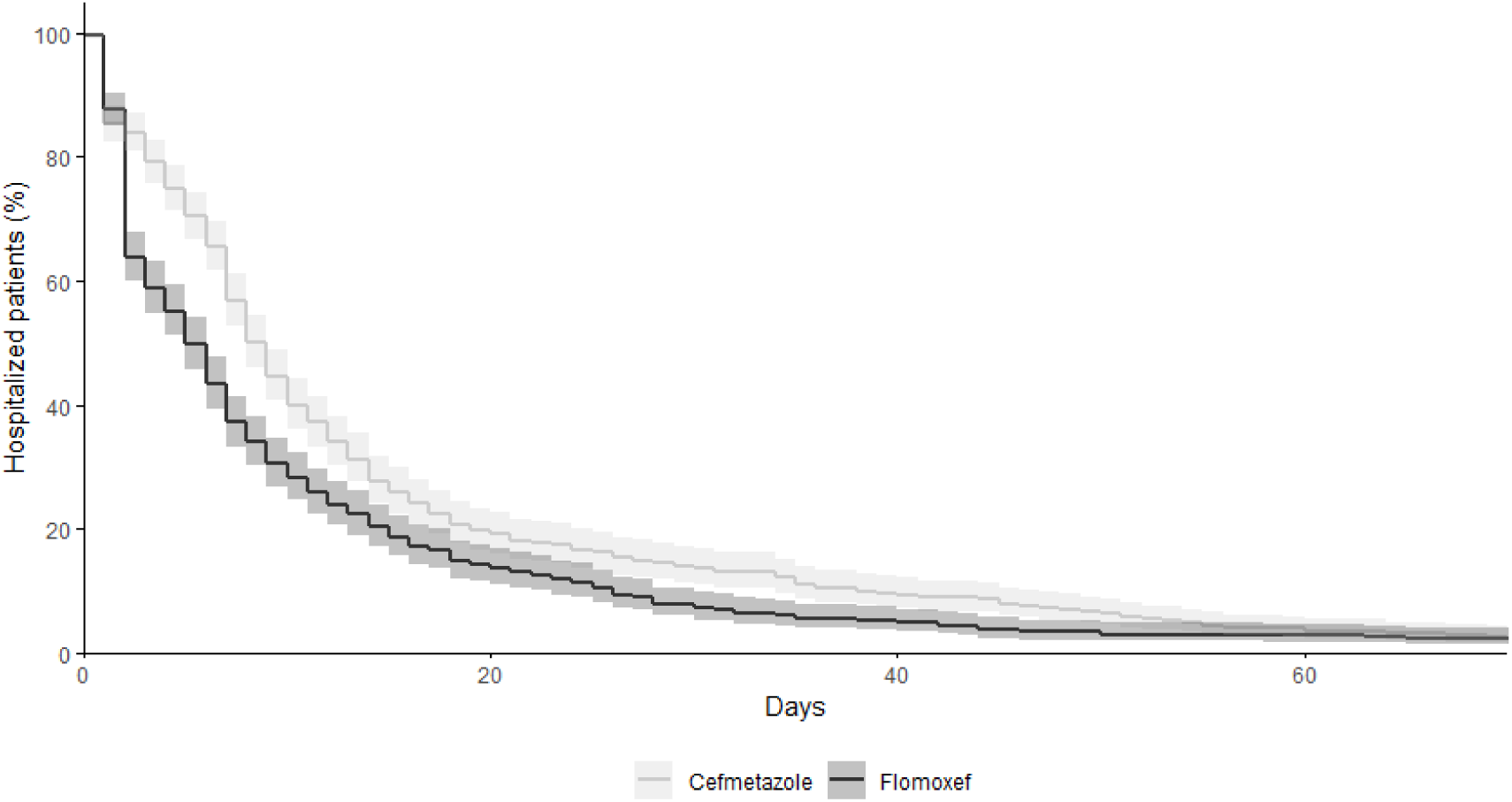
Kaplan–Meier plot illustrating survival post-discharge in Dataset 3. Kaplan-Meier curves for the length of hospital stay by an antimicrobial agent. Kaplan-Meier plots show hospitalization rates for patients in the cefmetazole and flomoxef groups from index date to survival discharge for Dataset 3.

### Secondary effectiveness outcome

The 28-day mortality rate was 2.71% (130/4796) in the cefmetazole group and 1.34% (9/673) in the flomoxef group, and multiple logistic regression analysis showed no significant difference between the two groups (OR: 0.89, 95% CI: 0.44–1.83, P = 0.760). Prescriptions for carbapenems were ordered for 6.01% (288/4796) and 4.61% (31/673) in the cefmetazole and flomoxef groups, respectively (OR: 0.92, 95% CI: 0.62–1.38, P = 0.698) (Table 3).

**Table 3.**
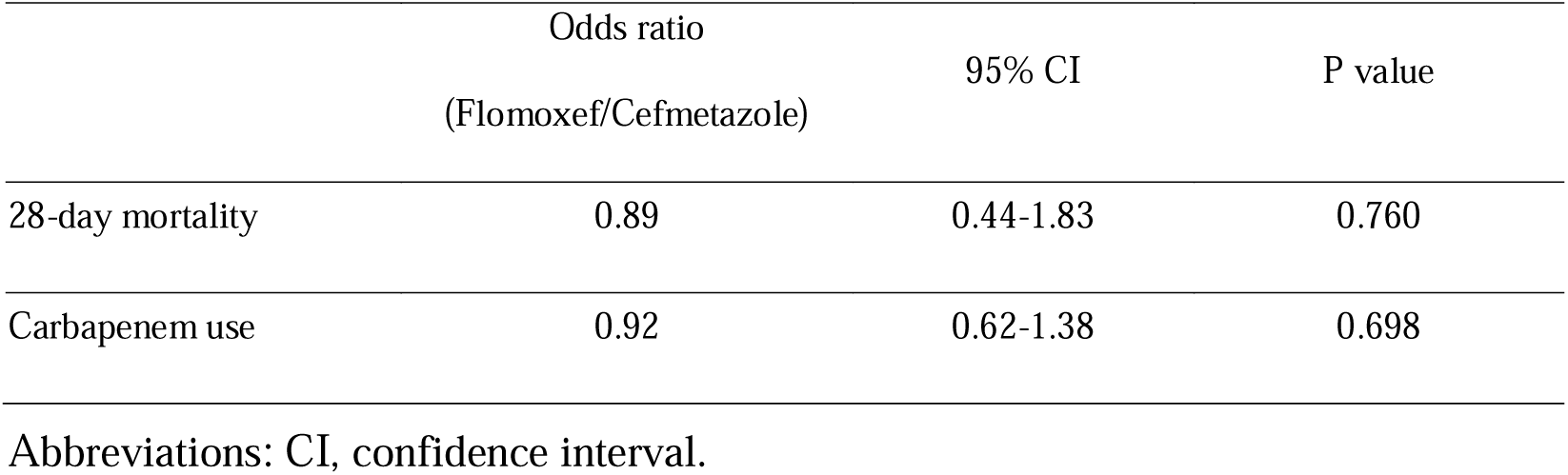
Secondary Efficacy Endpoints.

|  | Odds ratio<br>(Flomoxef/Cefmetazole) | 95% CI | P value |
| --- | --- | --- | --- |
| 28-day mortality | 0.89 | 0.44-1.83 | 0.760 |
| Carbapenem use | 0.92 | 0.62-1.38 | 0.698 |

Abbreviations: CI, confidence interval.

### Safety outcome

The incidences of adverse events related to the gastrointestinal, renal, and hepatic systems and hypersensitivity reactions were analyzed (Table 4). Renal failure was the most commonly reported adverse event in the cefmetazole group, affecting 7.5% of patients. This was followed by hepatic dysfunction, occurring in 2.3% of cases. Furthermore, gastrointestinal adverse events included *C. difficile* infection in 1.3% of patients in the cefmetazole group. In the flomoxef group, renal failure (3.6%) was also the most common, followed by hepatic dysfunction (2.5%) and other kidney dysfunction (0.9%). Kidney failure (P=0.04) and *C. difficile* infection (P<0.01) were significantly less common in the flomoxef group than in the cefmetazole group.

**Table 4.**
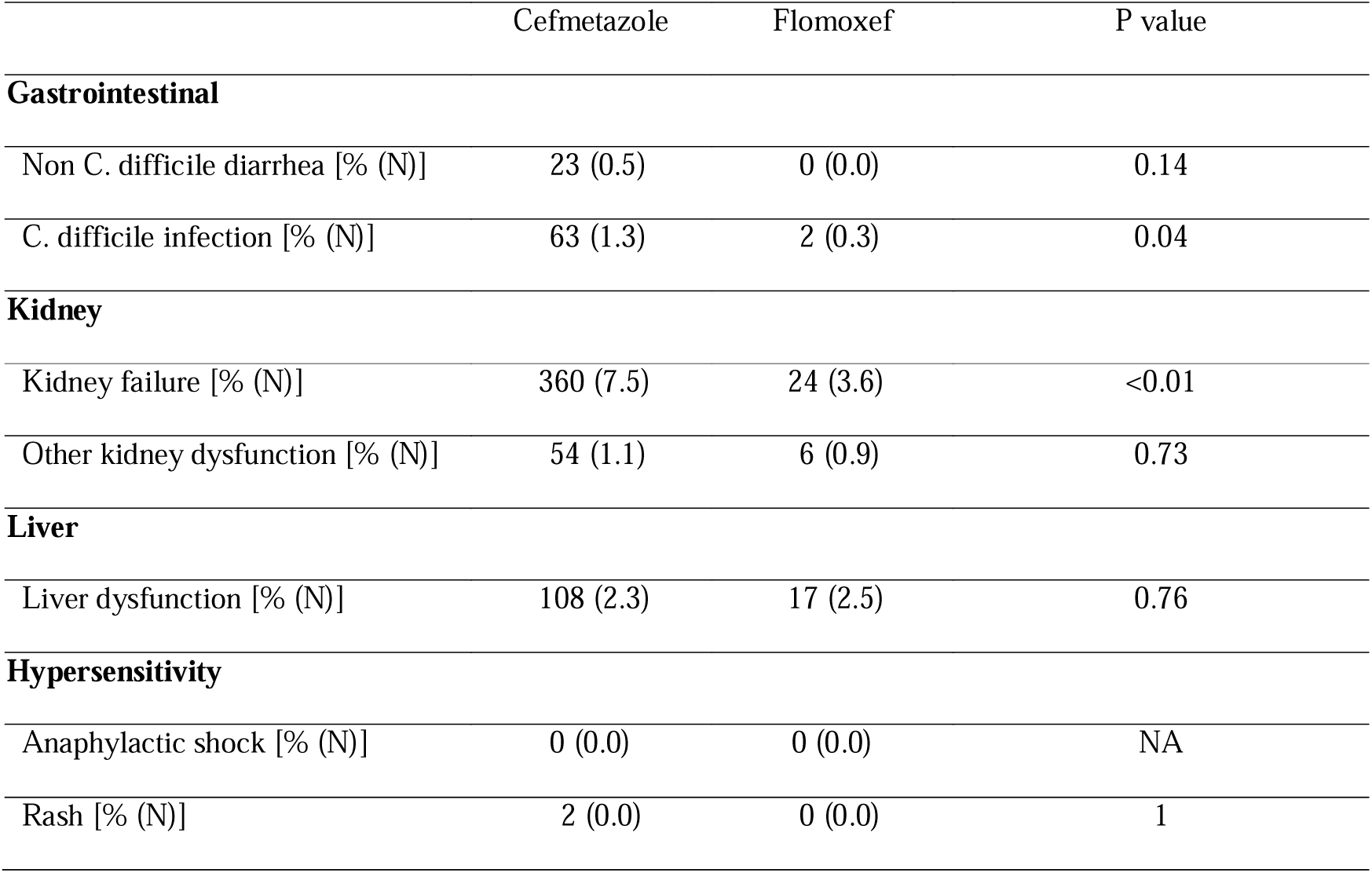
Safety Endpoints.

## Discussion

This study investigated the effectiveness and safety of flomoxef compared with cefmetazole in patients with UTIs. Our analysis of the Japanese surveillance data showed that the in vitro resistance rates of third-generation cephalosporin-resistant *E. coli* and *K. pneumoniae* for flomoxef were lower compared with cefmetazole. The *in vitro* evaluation of the combinational antimicrobial activities against an ESBL-producing multidrug-resistant strain demonstrated that both drugs exhibited similar patterns of interaction. Analysis of claims data revealed that the length of hospital stay for patients with UTI that required treatment were shorter in the flomoxef group than in the cefmetazole group, with no difference in 28-day mortality or the rate of additional carbapenem prescriptions. The incidence rates of kidney failure and *C. difficile* infection after therapy were significantly lower in the flomoxef group than in the cefmetazole group. Considering these data, flomoxef may be a potential treatment option for UTIs, including those caused by ESBL-producing bacteria, in clinical practice.

To date, no adequately powered randomized controlled trials have compared the relative efficacy of cephamycin and oxacephem against infections due to ESBL-producing bacteria. Several observational studies have compared the effectiveness of a cephamycin or an oxacephem, but all had relatively small numbers of patients [28]. As a result, clinical data on the use of a cephamycin or an oxacephem for bacteria including ESBL-producing bacteria are scarce, and the difference in efficacy between cefmetazole and flomoxef is unknown.

The CLSI defines the MIC breakpoints for cefmetazole against Enterobacterales as ≤16 μg/mL: Susceptible, 32 μg/mL: Intermediate, ≥64 μg/mL Resistant [18]. The JANIS data showed that flomoxef tended to have a lower frequency of MIC ≥64 μg/mL against 3GC-R *E. coli* and *K. pneumoniae*, including ESBL-producing bacteria, than cefmetazole. Indeed, a previous study reported lower MIC_90_ values of flomoxef than cefmetazole against ESBL-producing *E. coli* and *K. pneumoniae* [29]. These data suggest that flomoxef may be a treatment option for ESBL-producing bacteria.

The JMDC claims database used in this study encompasses data from 860 facilities across Japan, which are deemed to be representative of routine medical practices [30]. A previous study showed that over 80% of UTIs were due to either *E. coli* or *K. pneumoniae* [31, 32], and this epidemiological information can be extrapolated to the JMDC claim data as well. In addition, antimicrobial susceptibility testing results for clinical isolates during the same period as the claim data analysis may reflect the resistance status in the clinical background of the claims data. Therefore, part of the effectiveness of flomoxef observed in the claims data analysis may be explained by the difference in the rate of resistance between cefmetazole and flomoxef.

This study has several limitations. First, biases might exist in the clinical decision-making regarding the choice between cefmetazole and flomoxef. While there is no definitive evidence favoring the effectiveness of cefmetazole over flomoxef, it was observed that in claim data using present study, cefmetazole is more frequently prescribed to patients in a more severe general condition. Second, the claims data used in this study were limited to the hospitalization of interest, and any adverse events occurring after discharge from the hospital were not captured. The median length of hospital stay for flomoxef was about half that for cefmetazole, which may have resulted in a shorter observation period for adverse events in the flomoxef group.

As we have demonstrated the efficacy of flomoxef compared to cefmetazole in this study, the importance of flomoxef in the treatment of infections caused by ESBL-producing bacteria will increase internationally. The recent global neonatal sepsis observational cohort study led by GARDP has shown that susceptibility rates for flomoxef (71%) are comparable to those observed for meropenem (75%) in a subset of 108 *E. coli* and *K. pneumoniae* isolates. [31] Furthermore, retrospective multicenter studies in Japan have reported that treatment with cefmetazole or flomoxef for bacteremia caused by ESBL-producing *E. coli* is as effective as treatment with carbapenems [32]. Considering the recent shortage of important antimicrobials, especially beta-lactams, in clinical settings, flomoxef should be considered as an effective option against ESBL-producing bacteria, similar to cefmetazole and carbapenems. [33, 34]

## Acknowledgments

We would like to thank Yohei Doi for the critical reading of the manuscript. We are grateful to all the hospitals that participated in and contributed data to the JANIS.

## Author Contributions

Data collection, Analysis, Interpretation, and Statistical analysis: TN, TK, KY, KM, YM, MI, MS. Drafting manuscript: TN, TK, YM. Manuscript revision: TN, MG, SN, TK, KY, AH, YM, MS, KI. All the authors have read and approved the final version of the manuscript.

## Data availability statement

Data will be made available upon request from the corresponding author after the approval of the proposal.

