## Supplemental Table for "Clinical outcomes of flomoxef versus cefmetazole in hospitalized patients with urinary tract infections: Combined retrospective analyses of two real-world databases and *in vitro* data"

Supplementary Table 1. Definition of UTI

| Disease name | Follow-up Duration | ICD-10 Diagnosis Codes |
|---|---|---|
| **Study population** | | |
| Urinary tract infection | Index date | N39.0 Urinary tract infection, site not specified |

Abbreviations: ICD, International Statistical Classification of Diseases and Related Health Problems.

Supplementary Table 2. Definition of complicating disease

| Adverse Drug Event | Follow-up Duration | ICD-10 Diagnosis Codes |
|---|---|---|
| Sepsis | Index date | A40 Streptococcal sepsis<br>A41 Other sepsis |
| Shock | Index date | R57 Shock, not elsewhere classified |
| Respiratory failure | Index date | J96.0 Acute respiratory failure<br>J96.9 Respiratory failure, unspecified |

Abbreviations: ICD, International Statistical Classification of Diseases and Related Health Problems.

Supplementary Table 3. Definition of medical history

| **Disease name** | **Follow-up Duration** | **ICD-10 Diagnosis Codes** |
|---|---|---|
| diabetes mellitus | Before index date | E10-E14 Diabetes mellitus |
| urinary tract abnormalities | Before index date | N20-N23 Urolithiasis<br>N40 Hyperplasia of prostate<br>C64-C68 Malignant neoplasms of urinary tract<br>N35 Urethral stricture<br>C61 Malignant neoplasm of prostate |

Abbreviations: ICD, International Statistical Classification of Diseases and Related Health Problems.

Supplementary Table 4. Definition of safety outcomes adverse events

| Adverse Drug Event | Follow-up Duration | ICD-10 Diagnosis Codes |
|---|---|---|
| **Gastrointestinal** | | |
| Non-C. difficile diarrhea | Index date ~ 1 month | K52.3 Indeterminate colitis<br>K52.8 Other specified noninfective gastroenteritis and colitis<br>K52.9 Noninfective gastroenteritis and colitis, unspecified |
| C. difficile infection | Index date ~ 1 month | A04.7 Enterocolitis due to Clostridium difficile |
| **Kidney** | | |
| Kidney failure | Index date ~ 1 month | N17 Acute renal failure<br>N18 Chronic kidney disease<br>N19 Unspecified kidney failure |
| Other kidney dysfunction | Index date ~ 1 month | N28.9 Disorder of kidney and ureter, unspecified |
| **Liver** | | |
| Liver dysfunction | Index date ~ 1 month | K71.9 Toxic liver disease, unspecified<br>K76.9 Liver disease, unspecified |
| **Hypersensitivity** | | |
| Anaphylactic shock | Index date ~ 2 days | T78.2 Anaphylactic shock, unspecified<br>T88.6 Anaphylactic shock due to adverse effect of correct drug or medicament properly administered |
| Rash | Index date ~ 2 days | R21 Rash and other nonspecific skin eruption |

Abbreviations: ICD, International Statistical Classification of Diseases and Related Health Problems.
